# Profiling ACE2 expression in colon tissue of healthy adults and colorectal cancer patients by single-cell transcriptome analysis

**DOI:** 10.1101/2020.02.15.20023457

**Authors:** Haoyan Chen, Baoqin Xuan, Yuqing Yan, Xiaoqiang Zhu, Chaoqin Shen, Gang Zhao, Linhua Ji, Danhua Xu, Hua Xiong, TaChung Yu, Xiaobo Li, Qiang Liu, Yingxuan Chen, Yun Cui, Jie Hong, Jing-Yuan Fang

## Abstract

A newly identified novel coronavirus (2019-nCoV) has caused numerous acute respiratory syndrome cases in Wuhan China from December 2019 to Feb 2020. Its fast spreading to other provinces in China and overseas is very likely causing a pandemic. Since the novel coronavirus has been reported to be capable of endangering thousands of lives, it is extremely important to find out how the coronavirus is transmitted in human organs. Apart from fever and respiratory complications, gastrointestinal symptoms are observed in some patients with 2019-nCoV but the significance remains undetermined. The cell receptor angiotensin covering enzyme II (ACE2), which is the major receptor of SARS-nCoV, has been reported to be a cellular entry receptor of 2019-nCoV as well. Here, to more precisely explore the potential pathogen transmission route of the 2019-nCoV infections in the gastrointestinal tract, we analyzed the ACE2 RNA expression profile in the colon tissue of healthy adults and colorectal cancer patients of our cohort and other databases. The data indicates that ACE2 is mainly expressed in epithelial cells of the colon. The expression of ACE2 is gradually increased from healthy control, adenoma to colorectal cancer patients in our cohort as well as in the external Asian datasets. According to the expression profile of ACE2 in colon epithelial cells, we speculate adenoma and colorectal cancer patients are more likely to be infected with 2019-nCoV than healthy people. Our data may provide a theoretical basis for the classification and management of future 2019-nCoV susceptibility people in clinical application.

## Introduction

As one member of the family coronaviride, coronavirus are enveloped with non-segmented positive sense RNA, which are broadly distributed in humans and other mammals(*1*). Most of the coronavirus are harmless to human, except the beta coronaviruses, severe acute respiratory syndrome coronavirus (SARS-CoV)(*2, 3*) and Middle East respiratory syndrome coronavirus (MERS-CoV)(*4, 5*), which lead to numerous lethal respiratory disease cases in the past two decades. In December 2019, a series of unidentified pneumonia disease outbreaks was reported in Wuhan, Hubei province, China. In a subsequent pathogen identification process, 2019-nCoV (2019 novel coronavirus) was identified from the bronchoalveolar lavage fluid of these pneumonia disease patients(*6, 7*). This virus was identified as the cause of the deadly pneumonia disease in human (*7*). As of Feb 11, 2020, China has reported more than 42747 confirmed and more than 21675 suspected cases of 2019-nCoV infections across 33 Chinese provinces or municipalities, with 1017 fatalities. In addition, since 2019-nCoV is a novel coronavirus for human, there is no specifically effective drug for the infected patients.

Recently, Huang et al. reported that one diarrhea case was found in 38 patients with 2019-nCoV infections(*8*). In addition, some patients with diarrhea were constantly diagnosed in subsequent newly discovered 2019-nCoV infected cases, which indicate that 2019-nCoV may infect people via digestive systems. Although other groups have shown ACE2 expression profile in digestive system and lung by single-cell transcriptomes bioinformatics analysis from external datasets(*9, 10*), unfortunately, most data of these datasets are not from Asian adults. Therefore, to provide more implications of viral transmission in clinical diagnosis and treatment, it’s urgent to explore ACE2 expression in the digestive system of Asian people.

## Results

To investigate the ACE2 expression pattern in the gastrointestinal tract of Asian adult samples, we incorporated two public single cell RNA-seq data of colorectal cancer (CRC) cohorts from GEO (GSE97693 and GSE81861) and one cohort from Renji Hospital. We performed single cell analysis on 504 cells from GSE97693, including 485 cell from CRC (primary tumor) and 19 cells from healthy normal control (NC). We observed that ACE2 is expressed in 38.5%(194/504) of all human colon cells. We also observed that the ACE2 expressed cells were significantly enriched in tumors when compared with healthy normal tissues (39.6% vs 10.5%, p <0.05) (Fig. 1 A-C). Consistently, in GSE81861 dataset, ACE2 is expressed in 10.5%(62/590) of all human colon cells. We also observed that the ACE2 expressed cells were significantly enriched in colorectal tumors when compared with healthy normal tissues (12.5% vs 7.0%, p < 0.05) (Fig.1D-F). Those results intrigue us to investigated whether ACE2 is also enriched in precancerous lesion of colorectal tumor, such as colorectal adenoma(CRA). We performed single cell RNA profiling on the tissues from colorectal cancer, colorectal adenoma and healthy normal control from Renji Hospital hospital. Totally, 21,686 cells (4784 from CRC, 7130 from CRA and 9772 from NC samples) were collected after single cell RNA-seq quality control (Fig.2A). The ACE2 expressed cells were significantly enriched in CRC samples compared with CRA and NC samples (5.15% vs 2.94% and 1.75%, p<0.05) (Fig.2 B, C). In consistent with previous studies(*9, 10*), we also observed that ACE2 cell were mainly expressed in epithelial cells which highly express EPCAM(Fig.2D).

**Fig. 1.**
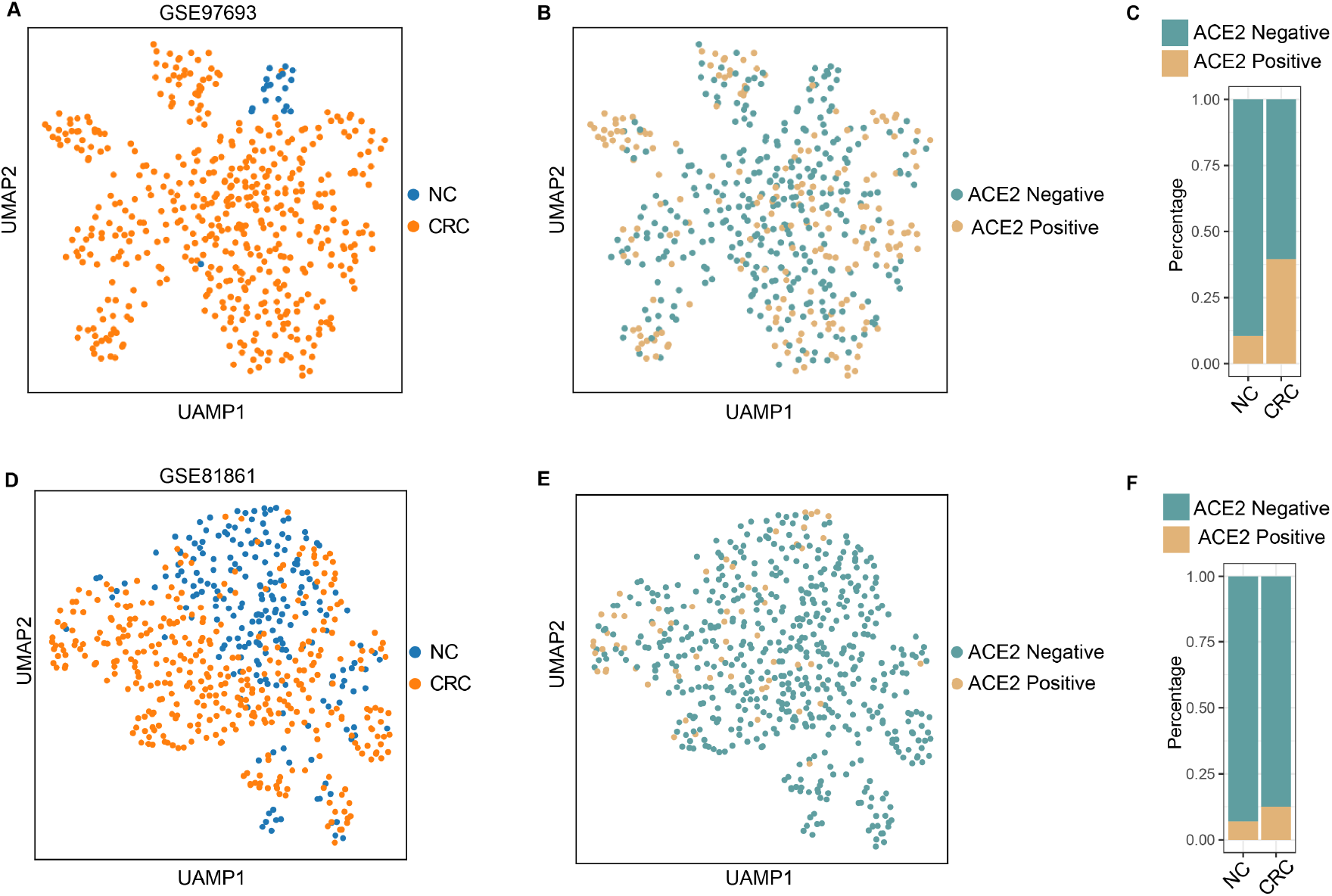
Single-cell analysis of colorectal cancer cells from public datasets (GSE97693 and GSE81861). (**A**). UMAP plots showing the distribution of CRC and NC cells in GSE97693. (**B**). UMAP plots showing the expression of ACE2 in GSE97693. (**C**). Stacked barplot for the expression of ACE2 in CRC and NC cell of GSE97693. (**D**). UMAP plots showing the distribution of CRC and NC in GSE81861. (**E**). UMAP plots showing the expression of ACE2 in GSE81861. (**F**). Stacked barplot for the expression of ACE2 in CRC and NC cell of GSE81861. Abbreviations: CRC, colorectal cancer; NC, healthy normal control; UMAP, Uniform Manifold Approximation and Projection

**Fig. 2.**
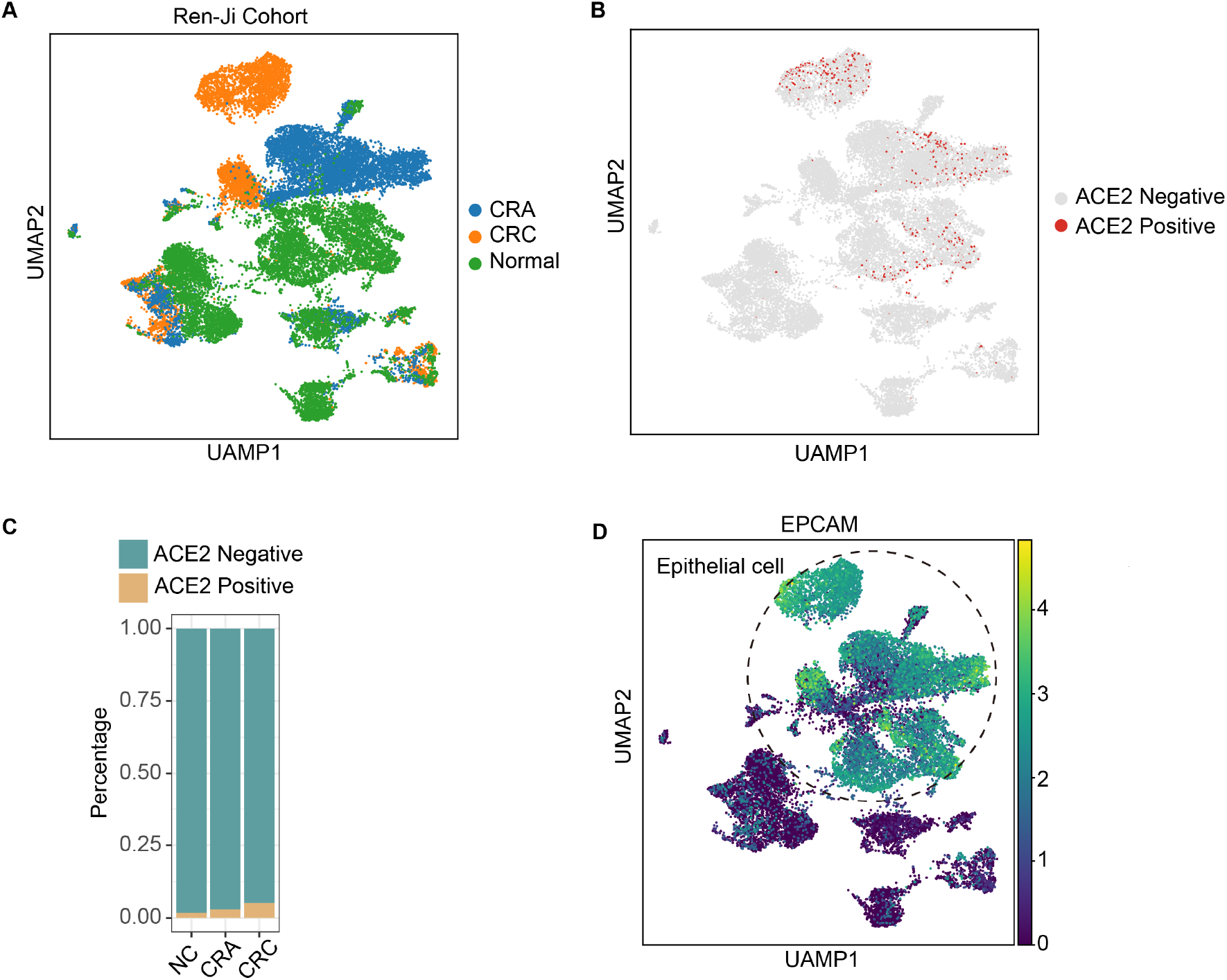
Single-cell analysis of colorectal cancer cells from Renji Cohort. (**A**). UMAP plots showing the distribution CRC, CRA and NC cells. (B). UMAP plots showing the expression of ACE2 in CRC, CRA and NC cells. (C). Stacked barplot for the expression of ACE2 in CRC, CRA and NC cells. (**D**). UMAP plots showing the expression of EPCAM (epithelial cell marker) in CRC, CRA and NC cells. Abbreviations: CRC, colorectal cancer; CRA, colorectal adenoma; NC, healthy normal control; UMAP, Uniform Manifold Approximation and Projection

## Methods

### Single cell sequencing

Single cell libraries were generated using the Chromium Single Cell 3′ library and gel bead kit v3 from 10x Genomics. Libraries were sequenced on the NovaSeq 6000 System (Illumina) at Shanghai GeneGenery BioTech Co.Ltd with 150 bp paired-end sequencing of reads. Gene counts were obtained by aligning reads to the hg38 genome (GRCh38) using CellRanger toolkit (version 3.0.2, 10X Genomics). The unique molecular identifier (UMI) count matrix was converted to anndata objects (version 0.6.22) using the Scanpy package v1.4.4 (pmid: 29409532). We utilized the following procedure to control for data quality: cells with fewer than 200 genes, and genes that are detected in less than 3 cells. Following the quality control procedure, the dataset consisted of 21,686 cells expressing 22,760 genes. The filtered gene expression matrix was normalized using Scanpy’s sc.pp.normalize_total function. A single-cell neighborhood graph was computed on the 50 first principal components that sufficiently explain the variation in the data using 15 nearest neighbors. Uniform Manifold Approximation and Projection (UMAP) was run for visualization. Cell types were annotated based on the expression of known marker genes.

External data sources: GSE97693 and GSE81861 were used for bioinformatics analysis. The reads or fragments per kilobase per million mapped reads (RPKM or FPKM) or transcripts per million mapped reads (TPM) were retrieved from GEO database(https://www.ncbi.nlm.nih.gov/geo/). In GSE97693 dataset, only TPM data were used for analysis.

## Discussion

The upper respiratory, gastrointestinal and central nervous system are usually the infection targets of coronavirus in humans and other mammals(*11*). A distinct coronavirus SARS-CoV was identified as the etiological agent of severe acute respiratory syndrome (SARS)(*2, 12, 13*). This virus may destroy the upper respiratory tract and damage the patient’s intestines by causing diarrhea(*14-16*). In December 2019, 2019-nCoV from the same family as SARS virus has been identified, and this virus can also cause fever, lower respiratory tract infection and gastrointestinal symptoms (*17*). The genome sequences of 2019-nCoV have been obtained by next-generation sequencing from pneumonia disease patients’ samples, and the spike protein of this virus was identified as well(*17*). This virus may directly bind the angiotensin converting enzyme II (ACE2), which was known as cell receptor for SARS-CoV (*6*). In addition, 2019-nCoV is able to enter lots of mammal cells with ACE2 receptor expression, but not cells without ACE2(*6*). Interestingly, 2019n-CoV just specifically uses ACE2 to infect mammal cells, but not other coronavirus receptor(*6*). Therefore ACE2 receptor may be the key component for 2019-nCoV to infect human and transmit in the host body.

It is reported that ACE2 is highly expressed in the lung and other digestive tract organs in public external databases(*9, 10, 18*). However, Asian adult data is rare in these datasets. Here, we firstly proved that ACE2 is expressed in the colon epithelial cells of Chinese adults. Furthermore, ACE2 expression is gradually increased from normal colon epithelium, adenoma to colorectal cancer patients’ tissues. Based on the expression of ACE2, colorectal cancer and adenoma patients are more likely to be infected and injured by 2019-nCoV, than healthy people.

As an angiotensin-converting enzyme (ACE) homologue(*19, 20*), ACE2 is a zinc metallopeptidase that catalyzes the conversion of angiotensin I (Ang I) and angiotensin II (Ang II) to angiotensin (1–9) and angiotensin (1–7), respectively(*21*).In addition to being a functional receptor for SARS-virus to enter mammal cells, ACE2 contributes substantially to the efficiency of SARS-CoV replication as well(*22*). Recently, 2019-nCoV was confirmed to share the same cell entry receptor, ACE2, as SARS-CoV(*6*). In addition, J. Byrnes et al. found that inhibition of ACE2 with GL1001, which is a selective inhibitor of this enzyme significantly reduced DSS-induced distal colon pathology, indicating that ACE2 may mediate the mucosal inflammation in DSS-induced inflammatory bowl disease (IBD) of mouse(*23*). Inhibition of ACE2 may increase the level of its substrates, such as gherlin and further block the production of pro-inflammatory cytokines(*24*). In a recent clinical report, 2019-nCoV may cause cytokine storm and multi-organ failure in severe pneumonia patients(*8*). Therefore, we speculates that 2019-nCoV may interact with ACE2 receptor in gastrointestinal tract, then further impair the intestinal mucous membrane barrier and increase the inflammatory cytokines production. With the intensification infection of 2019-nCoV, more inflammatory factors are produced, which eventually leads to a storm of inflammatory factors. Therefore, it is possible to use ACE2 inhibitors to treat 2019-nCoV patients by blocking ACE2 receptor of the intestinal epithelial cells in future clinical application.

In a word, our data support the intestinal tropism of the 2019-nCoV has some implications on clinical presentation and viral transmission. ACE2 expression is much higher in epithelia cells of colorectal cancer tissues than health control, which indicates that 2019-nCoV may be more harmful to colorectal cancer patients than health control people. Understanding of the tissue tropism of 2019-CoV on intestinal cells may also help to elucidate the pathogenetic mechanism of this virus and possibly help in the development of novel antiviral therapy. Future studies have to elucidate whether the replication of 2019-nCoV is depend on ACE2 in host cells, and whether 2019-nCoV binding to a co-receptor in addition to ACE2 might be involved in the specific infection of lung, gastrointestinal tract and other organs.

## Data Availability

All data and data processing scripts are available from the corresponding author upon request.

## AKNOWLEDGEMENTS

This project was supported in part by grants from the National Natural Science Foundation of China (81421001, 81871901,81874159).

## Reference

1. S. Su et al., Epidemiology, Genetic Recombination, and Pathogenesis of Coronaviruses. Trends Microbiol 24, 490–502 (2016).

2. T. G. Ksiazek et al., A novel coronavirus associated with severe acute respiratory syndrome. N Engl J Med 348, 1953–1966 (2003).

3. T. Kuiken et al., Newly discovered coronavirus as the primary cause of severe acute respiratory syndrome. Lancet 362, 263–270 (2003).

4. R. J. de Groot et al., Middle East respiratory syndrome coronavirus (MERS-CoV): announcement of the Coronavirus Study Group. J Virol 87, 7790–7792 (2013).

5. A. M. Zaki, S. van Boheemen, T. M. Bestebroer, A. D. Osterhaus, R. A. Fouchier, Isolation of a novel coronavirus from a man with pneumonia in Saudi Arabia. N Engl J Med 367, 1814–1820 (2012).

6. P. Zhou et al., A pneumonia outbreak associated with a new coronavirus of probable bat origin. Nature, (2020).

7. N. Zhu et al., A Novel Coronavirus from Patients with Pneumonia in China, 2019. N Engl J Med, (2020).

8. C. Huang et al., Clinical features of patients infected with 2019 novel coronavirus in Wuhan, China. Lancet, (2020).

9. Z. K. Hao Zhang, Haiyi Gong, D. Xu, Jing Wang, Zifu Li, Xingang Cui, Jianru Xiao, Tong Meng, Wang Zhou, Jianmin Liu, Huji Xu, The digestive system is a potential route of 2019-nCov infection: a bioinformatics analysis based on single-cell transcriptomes. bioRxiv, (2020).

10. Z. Z. Yu Zhao, Yujia Wang, Yueqing Zhou, Yu Ma, Wei Zuo, Single-cell RNA expression profiling of ACE2, the putative receptor of Wuhan 2019-nCov. bioRxiv, (2020).

11. S. Perlman, J. Netland, Coronaviruses post-SARS: update on replication and pathogenesis. Nat Rev Microbiol 7, 439–450 (2009).

12. C. Drosten et al., Identification of a novel coronavirus in patients with severe acute respiratory syndrome. N Engl J Med 348, 1967–1976 (2003).

13. R. A. Fouchier et al., Aetiology: Koch’s postulates fulfilled for SARS virus. Nature 423, 240 (2003).

14. J. S. Peiris et al., Coronavirus as a possible cause of severe acute respiratory syndrome. Lancet 361, 1319–1325 (2003).

15. J. S. Peiris et al., Clinical progression and viral load in a community outbreak of coronavirus-associated SARS pneumonia: a prospective study. Lancet 361, 1767–1772 (2003).

16. W. K. Leung et al., Enteric involvement of severe acute respiratory syndrome-associated coronavirus infection. Gastroenterology 125, 1011–1017 (2003).

17. R. Lu et al., Genomic characterisation and epidemiology of 2019 novel coronavirus: implications for virus origins and receptor binding. Lancet, (2020).

18. S. Z. Jun Wang, Ming Liu, Zhiyao Zhao, Yiping Xu, Ping Wang, Meng Lin, Yanhui Xu, Bing Huang, Xiaoyu Zuo, Zhanghua Chen, Fan Bai, Jun Cui, Andrew M Lew, Jincun Zhao, Yan Zhang, Haibin Luo, Yuxia Zhang, ACE2 expression by colonic epithelial cells is associated with viral infection, immunity and energy metabolism. MedRxiv, (2020).

19. M. Donoghue et al., A novel angiotensin-converting enzyme-related carboxypeptidase (ACE2) converts angiotensin I to angiotensin 1-9. Circ Res 87, E1–9 (2000).

20. S. R. Tipnis et al., A human homolog of angiotensin-converting enzyme. Cloning and functional expression as a captopril-insensitive carboxypeptidase. J Biol Chem 275, 33238–33243 (2000).

21. D. W. Lambert, N. M. Hooper, A. J. Turner, Angiotensin-converting enzyme 2 and new insights into the renin-angiotensin system. Biochem Pharmacol 75, 781–786 (2008).

22. W. Li et al., Angiotensin-converting enzyme 2 is a functional receptor for the SARS coronavirus. Nature 426, 450–454 (2003).

23. J. J. Byrnes et al., Effects of the ACE2 inhibitor GL1001 on acute dextran sodium sulfate-induced colitis in mice. Inflamm Res 58, 819–827 (2009).

24. T. Waseem, M. Duxbury, H. Ito, S. W. Ashley, M. K. Robinson, Exogenous ghrelin modulates release of pro-inflammatory and anti-inflammatory cytokines in LPS-stimulated macrophages through distinct signaling pathways. Surgery 143, 334–342 (2008).

